# Ensilication preserves high-molecular weight native DNA for clinical long-read sequencing

**DOI:** 10.1101/2025.10.26.25338579

**Authors:** Alexis Ferrasse, Rodrigo Mendez, John E. Gorzynski, Chloe Reuter, Jennefer N. Carter, Undiagnosed Diseases Network, Jonathan A. Bernstein, Matthew T. Wheeler, James L. Banal, Euan A. Ashley

## Abstract

Long-read DNA sequencing detects genetic variants and epigenetic modifications simultaneously, but optimal preservation of molecular length and base modifications typically requires cold-chain infrastructure. This infrastructure dependence restricts genomics to well-resourced laboratories and limits field research. We show that ensilication, the encapsulation of DNA within silica matrices, preserves DNA integrity at ambient temperature equivalent to −80 °C freezing. Using Genome-in-a-Bottle (GIAB) reference samples, we show that ensilicated DNA maintains sequencing performance comparable to frozen samples, while exhibiting resistance to degradation during repeated handling and accelerated weathering conditions simulating decades of ambient storage.. In real world samples from patients, native long-read sequencing of ensilicated samples resolved a de novo variant in the segmentally duplicated GTF2I locus in one case and detected methylation episignatures diagnostic of KDM2A-related disorder in another. Ensilication removes cold-chain dependence for diagnostic-quality long-read sequencing, expanding access to molecular diagnostics in resource-limited settings.

## Introduction

Long-read DNA sequencing technologies have transformed our ability to detect genetic variants and epigenetic modifications simultaneously from native DNA molecules, enabling comprehensive analysis of complex genomic regions, structural variants, and regulatory states ^1^. These capabilities have proven particularly valuable for diagnosing rare genetic disorders, where critical diagnostic information inaccessible to short-read methods includes structural variants, short tandem repeats, mobile element insertions, variants in segmentally duplicated loci, and disease-specific methylation signatures^2–5^. However, the full potential of native DNA sequencing requires maintaining molecular length, sequence fidelity, and base modifications. This preservation challenge has become a critical bottleneck in genomics.

DNA degradation during storage or handling occurs through multiple pathways: mechanical fragmentation reduces molecular length, hydrolytic and oxidative damage introduce sequence errors, and loss of epigenetic modifications erases regulatory information. Each form of degradation compromises sequencing quality and downstream analysis. Currently, preserving molecular length and native base modifications for native long-read sequencing requires continuous low-temperature storage, creating infrastructure dependencies of this technology to well-resourced facilities with reliable power, backup systems, and specialized shipping capability^6^. This cold-chain requirement has become increasingly problematic as genomic medicine expands globally, limiting access to advanced diagnostics in resource-limited settings and complicating field-based research in infectious disease surveillance and biodiversity studies.

The challenges extend beyond infrastructure. Repeated freeze-thaw cycles during routine sample access progressively damage DNA through mechanical shearing and chemical degradation, particularly affecting the ultra-long sequences essential for resolving structural variants and complex genomic regions. While chemical fossilization approaches, such as silica-based encapsulation ^7,8^, have emerged as potential alternatives for DNA storage, it remains unknown whether these methods can protect DNA from mechanical degradation while maintaining methylation patterns. Moreover, whether the encapsulation and de-encapsulation processes themselves introduce sequence errors introduced via chemical modifications has not been evaluated extensively. This is particularly critical for native sequencing platforms, where both molecular length and base modifications directly impact sequencing performance and downstream analysis.

Here we demonstrate that ensilication, in which DNA is chemically fossilized through encapsulation within silica-based particles, preserves DNA integrity equivalent to ultra-low-temperature freezing across molecular length, sequence fidelity, and genome-wide methylation patterns. Using Genome-in-a-Bottle (GIAB) reference samples, we show that ensilicated DNA maintains sequencing performance comparable to frozen samples, while exhibiting resistance to degradation during repeated handling and accelerated weathering conditions simulating decades of ambient storage. We validate clinical utility by performing native long-read sequencing on ambient-preserved samples from patients with rare genetic disorders, resolving variants in segmentally duplicated regions and simultaneously detecting genetic variants alongside disease-specific methylation episignatures. By eliminating cold-chain infrastructure requirements while maintaining complete DNA integrity, this approach enables broader access to long-read sequencing for both clinical diagnostics and research applications in diverse global settings.

## Results and Discussion

### Ensilication preserves molecular length, sequence fidelity, and genome-wide methylation patterns equivalent to ultra-low temperature freezing

We systematically compared DNA preservation by ensilication to conventional −80 °C storage across molecular length, sequence fidelity, and epigenetic modifications. Using the gold-standard GIAB reference samples, we show this method preserves DNA quality comparable to ultra-low temperature storage while providing superior stability during routine handling.

We rigorously evaluated ensilication against conventional −80°C storage using three benchmark GIAB reference samples: HG002, HG003, and HG004. Following preservation via either method, samples were processed identically using Oxford Nanopore’s Native Ligation Sequencing Kit V14 and subjected to comprehensive quality assessment. The resulting multi-parameter analysis revealed high consistency between ensilication and freezing methods across all measured metrics. Read length density plots (**Fig. 1a**) demonstrated closely matching distributions between ensilicated and frozen samples across all three genomes. Cumulative base percentage analysis (**Fig. 1b)** further validated this observation, with N50 values showing limited variation between storage methods. K-mer frequency analysis (**Fig. 1c**) produced nearly identical logarithmic distributions between preservation methods, indicating that ensilication maintains consistent nucleotide sequence patterns without introducing detectable biases in the resulting sequencing reads. Quality score distributions (**Fig. 1d**) were notably congruent between ensilicated and frozen samples across all three genomes, with both methods yielding mean Q-scores primarily in the 17–20 range. Statistical analysis confirmed the equivalence of the two preservation methods. When comparing ensilicated versus frozen samples across all three genomes, we found no significant differences in any key sequencing metric, including N50 values (Mann-Whitney U test, p = 1.0), median read lengths (Mann-Whitney U test, p = 0.1), and quality scores (Mann-Whitney U test, p = 0.7) (**Table 1**).

**Table 1.** Summary of sequencing metrics for frozen and ensilicated DNA samples.

| Genome | Storage condition | N50 [bp] | Median length [bp] | Median Q-score |
| --- | --- | --- | --- | --- |
| HG002 | Frozen | 9729 | 1634 | 18.3 |
|  | Ensilicated | 9222 | 2452 | 18.7 |
| HG003 | Frozen | 10807 | 2028 | 18.6 |
|  | Ensilicated | 10179 | 2579 | 18.9 |
| HG004 | Frozen | 9129 | 2029 | 18.4 |
|  | Ensilicated | 8094 | 2068 | 18.1 |

**Figure 1.**
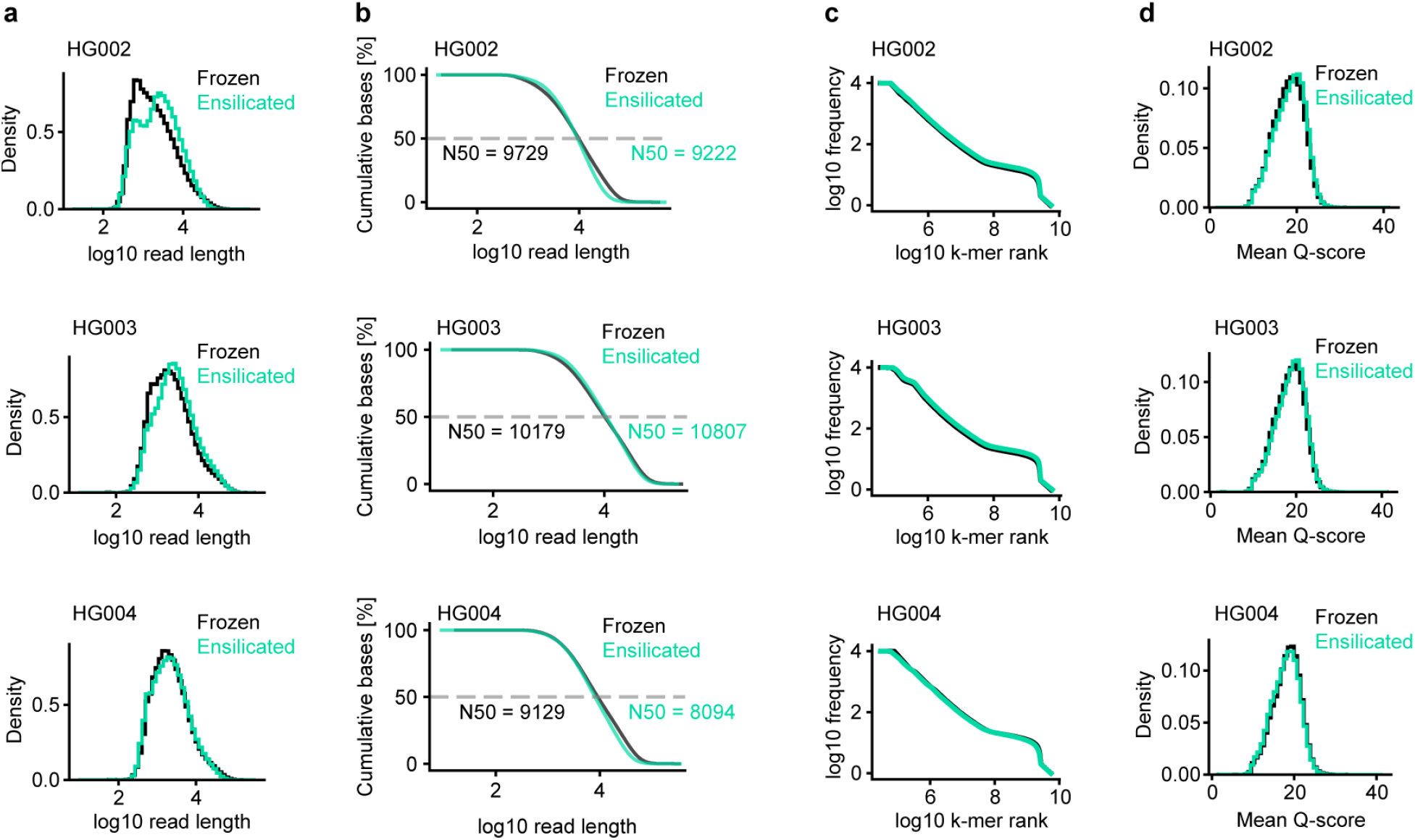
Ensilication maintains sequencing integrity comparable to conventional freezing across multiple quality metrics. **a**, Read length density distributions comparing frozen (black) and ensilicated (teal) DNA preparations across three distinct GIAB reference samples: HG002, HG003, and HG004. **b**, Cumulative base percentage plotted against read length, with N50 values indicated for both frozen and ensilicated conditions. **c**, Log-log k-mer rank frequency distributions comparing frozen and ensilicated samples. **d**, Mean Q-score density distributions demonstrating equivalent sequencing quality between frozen and ensilicated samples.

Variant calling analysis against GIAB benchmarks revealed high concordance for both SNP and small indel detection. SNP calling achieved consistently high precision, recall, and F1 scores (>0.95) across all three samples for both storage methods (**Fig. 2a**). While indel detection showed lower performance metrics relative to SNPs, this is characteristic of the sequencing technology rather than a reflection of storage conditions^9,10^. Importantly, the indel detection performance remained comparable between ensilicated and frozen samples, indicating that ensilication does not introduce additional challenges for variant calling. Structural variant (SV) analysis using Truvari^11^ demonstrated excellent concordance between storage methods, with precision, recall, F1 scores, and genotype concordance all exceeding 0.9 across all three samples (**Fig. 2b**). When benchmarking against the GIAB SV truth set for HG002, both storage methods showed comparable performance (**Fig. 2c**), confirming that ensilication maintains the ability to accurately detect complex genomic variants.

**Figure 2.**
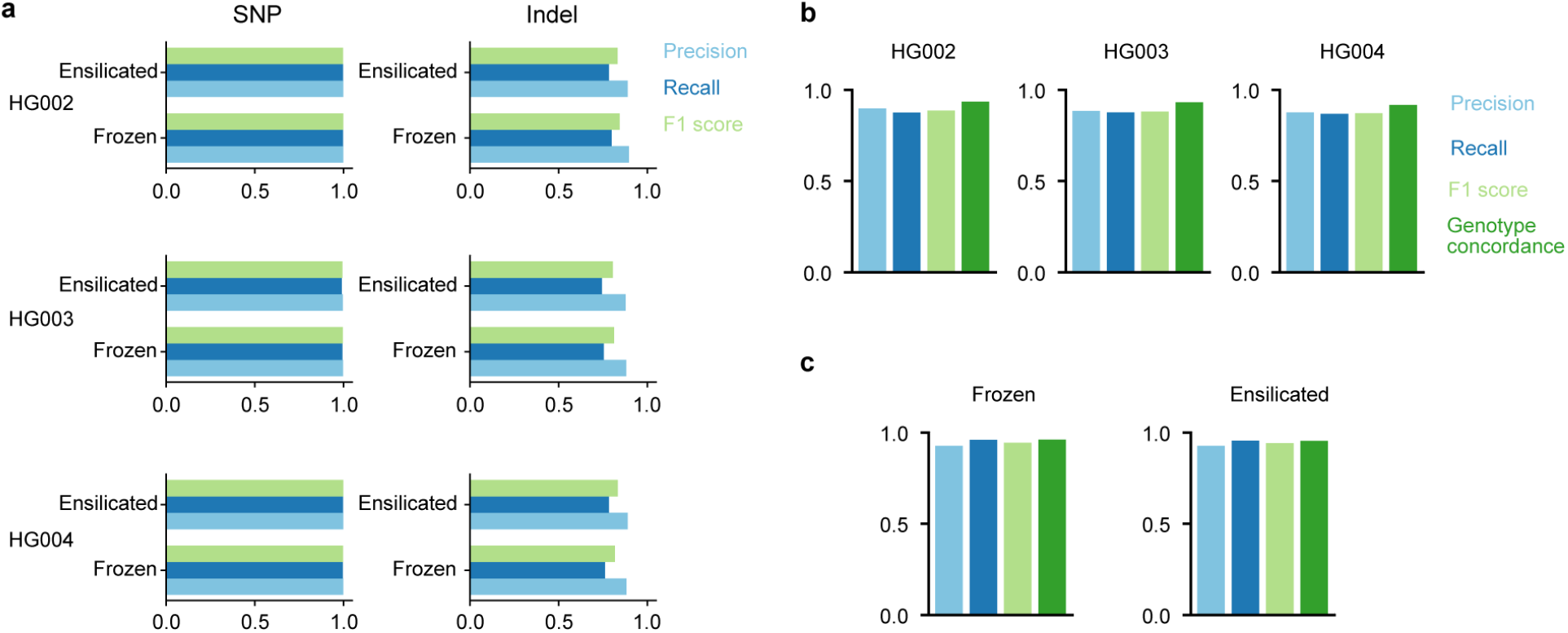
Ensilication preserves variant calling performance comparable to conventional freezing. **a**, Comparison of SNP (left) and indel (right) detection performance, measured by precision, recall, and F1 scores, for GIAB reference samples HG002, HG003, and HG004 stored by freezing or ensilication. **b**, SV detection comparison using frozen samples as the truth set, demonstrating consistently high precision, recall, F1 scores, and genotype concordance for ensilicated samples across all three genomes. **c**, Performance benchmarking of both ensilicated and frozen HG002 samples against the established GIAB structural variant truth set.

The ability to preserve both genetic and epigenetic information is crucial for comprehensive genomic analysis. CpG methylation, where a methyl group is added to cytosine nucleotides followed by guanine in the DNA sequence, is a key epigenetic modification that influences gene expression. At each CpG site, methylation levels (β values) range from 0% (completely unmethylated) to 100% (fully methylated). When comparing these β values between storage conditions using density scatter plots, we observed strong agreement across the full methylation spectrum: sites that were highly methylated in frozen samples remained highly methylated in ensilicated samples, and similarly for unmethylated sites. This concordance was quantified by Pearson correlation coefficients ranging from 0.93 to 0.95 across all three samples (**Fig. 3a**). However, while Pearson correlation quantifies the linear relationship between methylation levels, it does not directly measure how accurately ensilication preserves biologically meaningful methylation states. To specifically assess this, we performed receiver operating characteristic (ROC) analysis, using frozen samples as the reference standard. By binarizing methylation states into biologically relevant categories (methylated: β ≥ 50%, unmethylated: β < 50%) in the frozen samples, we evaluated the ability of ensilicated samples to correctly classify methylation at individual CpG sites. ROC analysis revealed high area under the curve (AUC) values (0.97–0.98), confirming that ensilication reliably maintains biologically relevant methylation classifications (**Fig. 3b**). Additionally, to further validate the accuracy and reliability of methylation measurements obtained from ONT sequencing, both ensilicated and frozen, we compared them to established whole-genome bisulfite sequencing (WGBS) methylation profiles derived from the HG002, HG003, and HG003 bedgraph data of the EpiQC study^12^. WGBS serves as a reference method, enabling us to assess the overall quality and consistency of ONT-based methylation data. This comparison demonstrated strong correlation (Pearson’s r ≥ 0.89), comparable to the previously reported correlation between ONT and EmSeq WGBS data in the EpiQC study^12^, thus confirming that ensilication does not compromise the reliability of epigenetic information (**Fig. 3c**). Collectively, these results demonstrate that ensilication effectively preserves DNA length, sequence integrity, and biologically relevant epigenetic information.

**Figure 3.**
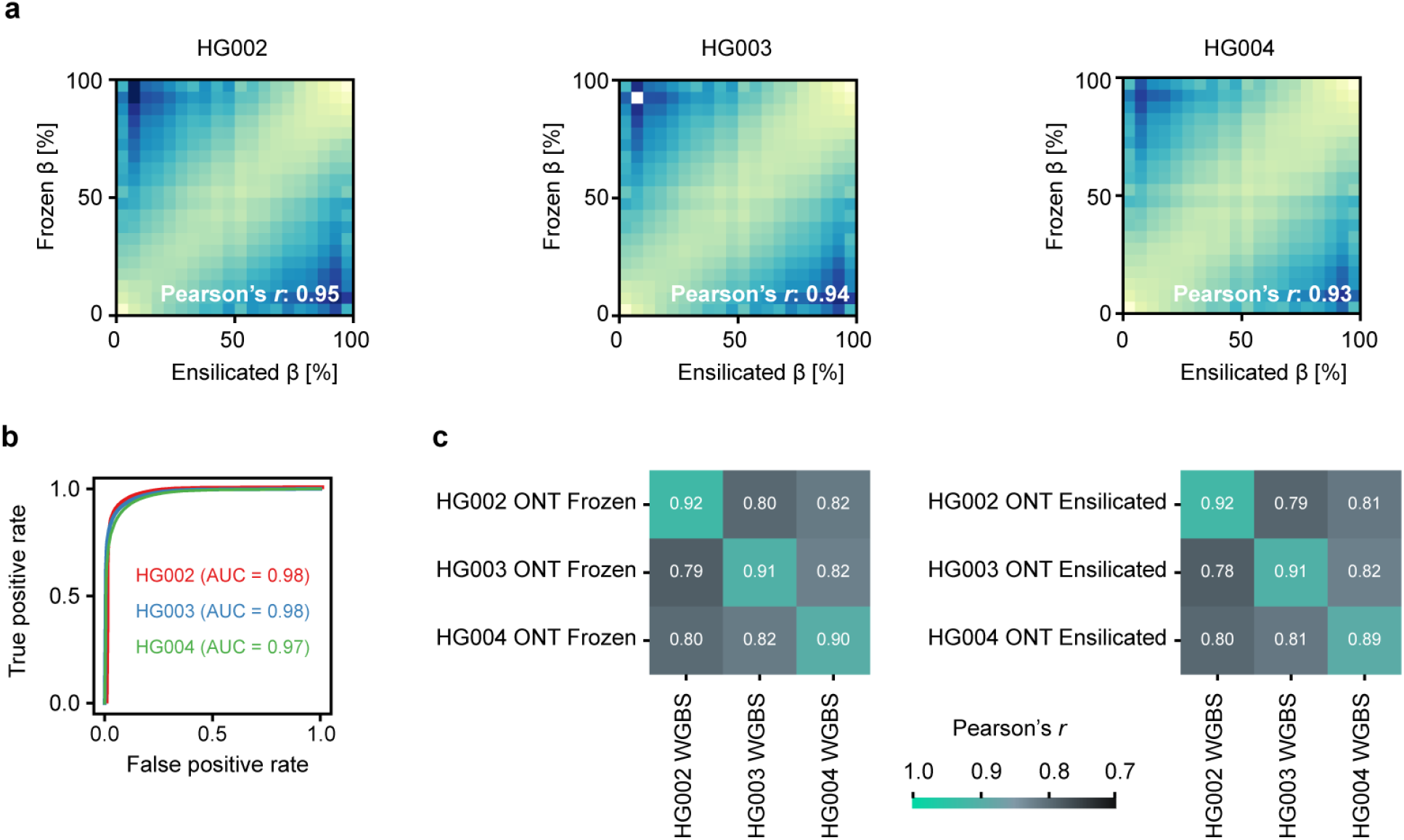
Ensilication preserves CpG methylation patterns comparable to conventional freezing. **a**, Density scatter plots comparing methylation levels (β values) between frozen and ensilicated samples for GIAB reference samples HG002, HG003, and HG004. **b**, ROC curves evaluating the accuracy of ensilicated samples in classifying biologically relevant methylation states, methylated vs. unmethylated, using frozen samples as ground truth. High AUC values confirm accurate preservation of methylation status by ensilication. **c**, Heatmaps showing Pearson correlation between ONT sequencing methylation profiles for frozen and ensilicated samples and EMSeq WGBS data from the EpiQC study, confirming that ensilication maintains methylation measurement quality comparable to standard benchmarks.

### Ensilication enables diagnosis in structurally complex loci and episignature profiling

Having established that ensilication maintains DNA quality equivalent to freezing across molecular length, sequence accuracy, and methylation patterns using reference samples, we next evaluated whether ambient-preserved DNA supports clinically meaningful analyses. GIAB samples provide gold-standard benchmarking but they represent idealized genomic contexts. Clinical diagnostics frequently require resolving variants in structurally complex regions where paralogs complicate read assignment, or interpreting variants of uncertain significance where epigenetic functional data provides critical evidence. To test whether ensilicated DNA preserves the information content necessary for such challenging diagnostic scenarios, we performed native long-read sequencing on ambient-preserved samples from patients with rare genetic disorders.

### Validation in a segmentally duplicated locus: de novo *GTF2I* variant in a proband with autism spectrum disorder

The proband is a male in his early twenties with autism spectrum disorder and intellectual disability. Developmental history was notable for language regression in early childhood. Current findings include absent speech, repetitive behaviors, sensory hypersensitivity, macrocephaly, and sleep disturbances (**Fig. 4a** and **4b**). Karyotype, Fragile X testing, and chromosomal microarray were unremarkable.

**Figure 4.**
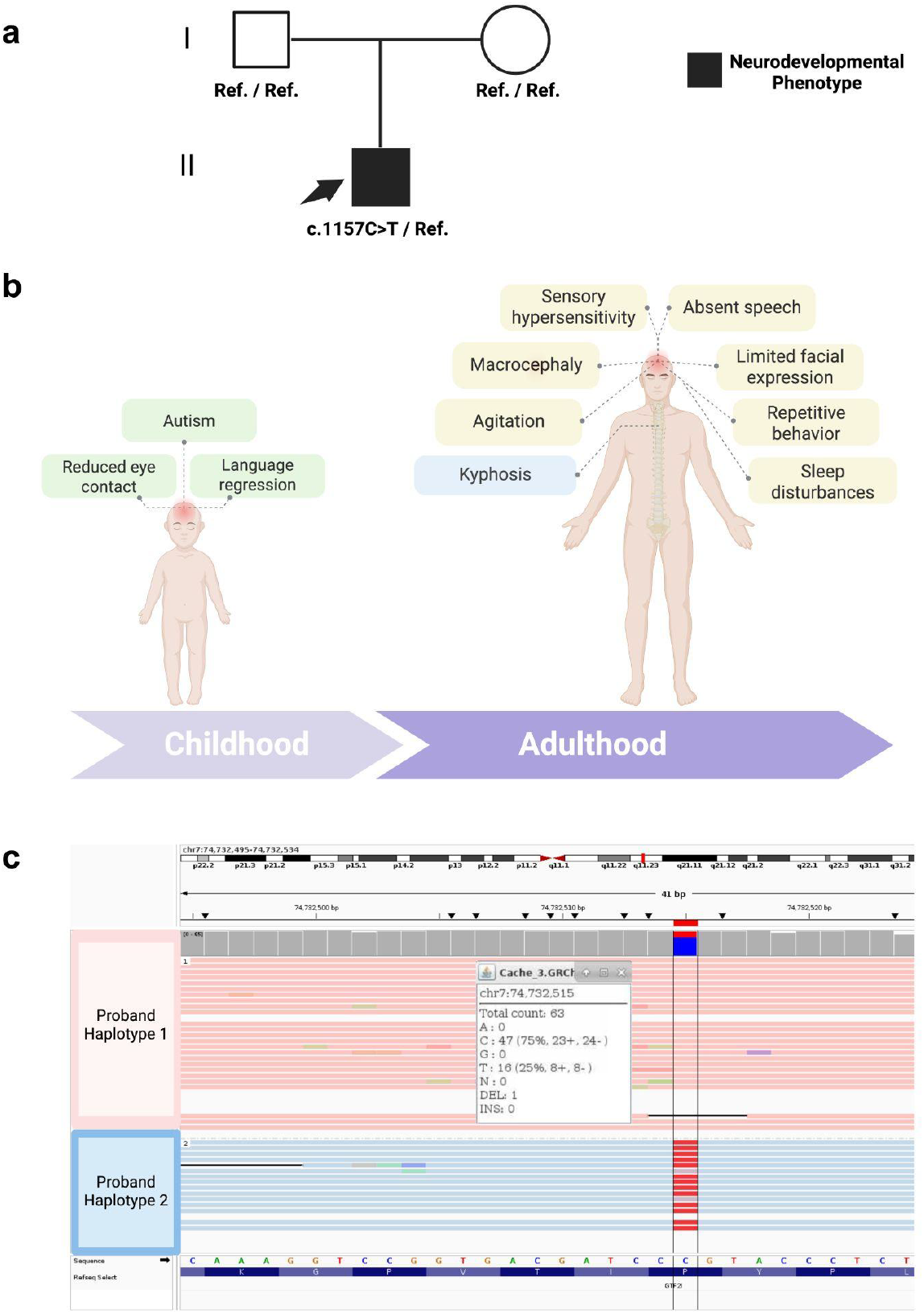
Clinical synopsis and phased long-read evidence for a heterozygous GTF2I variant. **a**, Pedigree with the proband indicated by the arrow. **b**, Lifespan phenotype summary. In childhood, the proband exhibited reduced eye contact, language regression, and autistic behavior. In adulthood, the proband shows absent speech, limited facial expression, sensory hypersensitivity, macrocephaly, agitation, repetitive behavior, sleep disturbances, and kyphosis. **c**, IGV view of ONT native-DNA reads aligned to GRCh38 at *GTF2I* (7q11.23; vertical line at chr7:74,732,515). Reads are partitioned by phase, with Proband Haplotype 1 (top) carrying the reference base and Proband Haplotype 2 (bottom) carrying the alternate base. Red mismatches mark the alternate allele, and the inset shows base counts. The phasing confirms a single-haplotype heterozygous SNV in GTF2I at a segmentally duplicated locus.

Clinical trio genome sequencing identified a de novo heterozygous missense variant in *GTF2I* (general transcription factor IIi, MIM^13^ 01679) (NM_032999.4:c.1157C>T; p.Pro386Leu; GRCh38 chr7:74,732,515 C>T), absent from gnomAD v4.1.0^14,15^ and predicted to be deleterious (CADD^16^ 28). This variant is listed in ClinVar^17^ as of uncertain significance (VCV003359229.2). *GTF2I* is located within the Williams-Beuren syndrome critical region (WBS, MIM^13^ 194050). Because *GTF2I* resides within the 7q11.23 segmental duplication block and has a closely related paralog/pseudogene (*GTF2IP1*), accurate read assignment is a known challenge^18^. To verify locus specificity, we used ONT long-read sequencing. ONT reads covering unique paralog-distinguishing motifs confirmed the variant’s position (read depth of 63×, 16 alternate reads (VAF 0.25)) in *GTF2I* (not *GTF2IP1*) (**Figure 4c**), with phasing consistent with heterozygosity and trio short-read genome data supporting a de novo origin. EpiSign^19^ methylation profiling results were inconclusive for the conditions tested. In future analyses of this case, we plan to evaluate ONT-derived methylation signals using newer long-read classifiers to enable quantitative episignature interpretation^20, 21^.

Recent evidence supports a monogenic GTF2I-related neurodevelopmental disorder^18^. A multicenter case series identified eight unrelated individuals with heterozygous *GTF2I* alterations, including de novo nonsense, splice-site, missense, in-frame deletion, and intragenic deletion variants, as well as one case with reduced *GTF2I* expression. Affected individuals shared global developmental delay or intellectual disability with frequent autistic features and facial dysmorphism partly reminiscent of Williams–Beuren syndrome. RNA sequencing confirmed aberrant splicing or decreased expression for several variants, consistent with a haploinsufficiency mechanism. The study also highlighted technical challenges due to homology with *GTF2IP1* and demonstrated that long-read sequencing can aid in variant confirmation. These findings provide human genetic validation that deleterious *GTF2I* variation can underlie isolated neurodevelopmental disease and align with prior functional models of *GTF2I* dosage sensitivity^22^.

### Genetic variant confirmation and episignature detection in a *KDM2A* case

The proband is a pre-adolescent female evaluated for dysmorphic features, generalized hypotonia, global developmental delay, feeding difficulties with early failure to thrive requiring nasogastric tube feedings, autism spectrum disorder, and attention-deficit/hyperactivity disorder with behavioral dysregulation. Prior chromosomal microarray, exome sequencing, and genome sequencing were nondiagnostic. Re-analysis identified a de novo heterozygous *KDM2A* (lysine demethylase 2A, MIM^13^ 605657) variant, NM_012308.3:c.1772T>C (p.Met591Thr), confirmed by Sanger sequencing. The clinical phenotype is consistent with *KDM2A*-related neurodevelopmental disorder as described in recent case series^23^, establishing its clinical relevance within a syndromic neurodevelopmental disorder. Using ONT long-read sequencing, we confirmed the variant’s position (read depth of 14×, 8 alternate reads (VAF 0.57)). We then used native DNA cytosine methylation calls from the same ONT dataset to visualize representative loci implicated in the *KDM2A* methylation signature and compared the proband qualitatively with ambient-preserved control specimens without *KDM2A* variants (**Fig. 5**). The proband exhibited hypermethylated CpG islands, including CYP26C1 CpG255, RB1 CpG85, and HS3ST3B1 CpG267. The qualitative pattern aligned with the *KDM2A*-positive cluster described in the cohort analysis^23^. Due to the limited scope of our ONT subset, these methylation findings are presented descriptively to demonstrate feasibility rather than to provide classifier-grade output. Future work could leverage emerging ONT-based methylation classifiers to enable quantitative classification^20, 21^.

**Figure 5.**
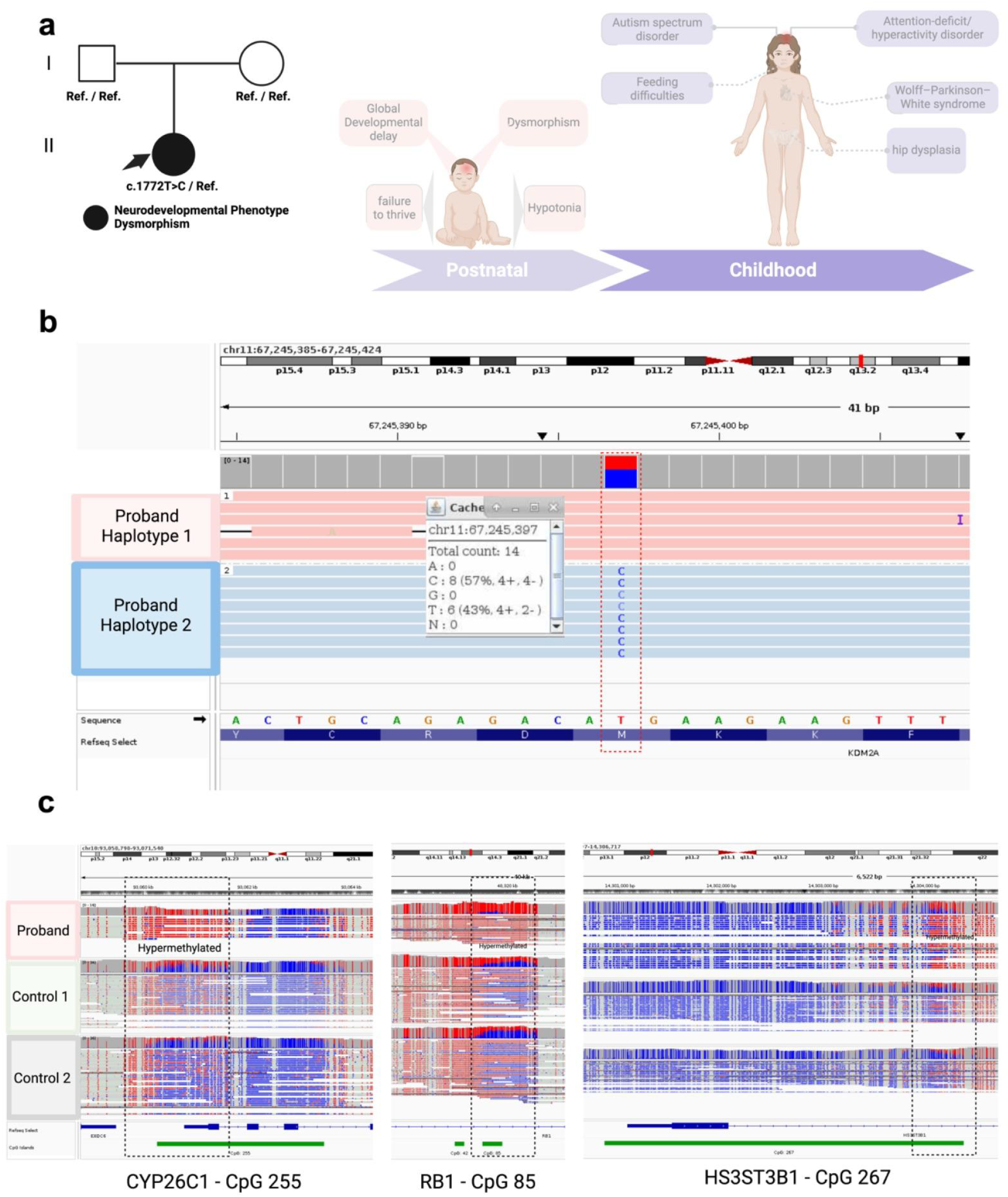
Clinical course, haplotype-resolved confirmation, and representative DNA-methylation CpG islands for a *KDM2A* variant. **a**, Pedigree and phenotype timeline. The proband (arrow, filled symbol) carries a de novo *KDM2A* c.1772T>C variant (parents Ref./Ref.) and presents with neurodevelopmental features consistent with *KDM2A*-related disorder. **b**, IGV view of Oxford Nanopore native-DNA reads aligned to GRCh38 at *KDM2A* (chr11:67,245,397). Reads are partitioned by phase, with Proband Haplotype 1 carrying the reference allele and Haplotype 2 carrying the alternate allele. Base counts support a heterozygous single-nucleotide variant present on a single haplotype. **c**, Representative native-DNA methylation tracks at three CpG sites used in the *KDM2A* episignature (CYP26C1 CpG255, RB1 CpG85, HS3ST3B1 CpG267). The proband exhibits focal hypermethylation relative to two controls (IGV convention: red, methylated; blue, unmethylated; dashed boxes indicate comparison windows), providing orthogonal functional support for the *KDM2A* variant.

## Conclusion and Outlook

While genomic sequencing is widely accessible globally, the cold-chain dependence for native long-read DNA sequencing has constrained access to its unique capabilities for patients with rare diseases and restricted research in settings where maintaining ultra-low temperature storage is impractical or impossible. By demonstrating that ensilication preserves DNA integrity equivalent to freezing across molecular length, sequence fidelity, and methylation patterns, we establish a foundation for infrastructure-independent genomics. Notably, preliminary stability studies suggest potential advantages over conventional freezing. Ensilicated samples showed less degradation than frozen samples after repeated handling cycles, reducing the need for aliquoting, and accelerated weathering experiments demonstrated maintained fragment size distributions compared to progressive degradation of dried DNA controls (**Supplementary Figure 1**). However, these findings require validation through long-term real-world storage studies across diverse environmental conditions before definitive conclusions about extended ambient stability can be drawn.

Ensilication enables ambient-temperature sample collection and shipping for native long-read sequencing. Samples collected at community hospitals or international clinics can be shipped at ambient temperature, undergo long-read sequencing weeks or months later, and still yield diagnostic-grade data including episignature analysis. Our *GTF2I* and *KDM2A* cases demonstrate this capability directly. Both samples were preserved at ambient temperature yet retained sufficient quality to resolve variants in segmentally duplicated regions and detect disease-specific methylation patterns.

Beyond access, ensilication addresses the hidden costs of cold storage: continuous energy consumption, carbon emissions from specialized shipping, and the progressive sample degradation that occurs through freeze-thaw cycles during routine laboratory use. As genomic medicine scales globally, transitioning from cold-dependent to ambient-stable preservation represents both an equity imperative and a sustainability opportunity. Critical questions remain for future investigation. Systematic long-term stability studies under varied temperature, humidity, and handling conditions will establish performance boundaries for ambient storage. Integration with point-of-collection workflows could enable decentralized sample processing, reducing time from collection to diagnosis. Extension to other sample types, tissue biopsies, circulating tumor DNA, microbial communities, would broaden utility across clinical and research applications. Additionally, formal cost-benefit analyses comparing cold-chain versus ambient preservation across different use cases will inform practical implementation strategies.

Cold-chain infrastructure has long been a practical requirement for preserving DNA suitable for advanced molecular diagnostics. Our demonstration that ensilication maintains DNA integrity at ambient temperature, equivalent to conventional freezing across molecular length, sequence fidelity, and methylation patterns, suggests this requirement may be relaxed for many applications. While systematic evaluation of long-term ambient stability under diverse real-world conditions remains necessary, the data presented here establish that diagnostic-quality preservation at ambient temperature is achievable. As sequencing platforms continue to become more portable and accessible, the geographic scope of genomic medicine need no longer be strictly defined by infrastructure limitations in sample preservation.

## Methods

### Study cohort

We evaluated two participants enrolled in the Undiagnosed Diseases Network (UDN) using Oxford Nanopore Technologies (ONT) long-read whole-genome sequencing (lrWGS) from whole-blood DNA: Case 1 with a candidate variant in GTF2I, and Case 2 with a research-solved candidate in KDM2A that we used as a positive control for our post-encapsulation workflow.

### Genomic DNA encapsulation and library preparation

Genomic DNA (3 µg) was dissolved in 1000 µL nuclease-free water (Thermo Fisher; catalog number: AM9916) in a 1.5 mL Eppendorf LoBind tube. Encapsulation particles from Cache DNA, Inc. (3 mg) were added, and the suspension was mixed by turning the tube upside down 3 times. Ensilication reagents from Cache DNA, Inc. were then added, and the mixture was shaken for 48 hours at 1600 rpm using a Bioshake iQ (Bulldog Bio; catalog number: 1808-0506).

The suspension was pelleted with a microcentrifuge for 10 seconds and the supernatant was carefully removed. The pellet was washed with 0.1% Tween-20 and stored for 30 days at ambient temperature until further processing. For DNA retrieval, 300 µL of de-encapsulation buffer (Cache DNA, Inc.) was added, and the mixture was incubated at ambient temperature for 5 minutes. De-encapsulated DNA was purified using a Multi-screen 96-well filter plate (Millipore Sigma; catalog number: MSNU03010). Briefly, 300 µL of the sample was added to a well and filtered to dryness. A volume of 150 µL of nuclease-free water was then added and also filtered to dryness. Finally, a volume of 55 µL of resuspension buffer (20 mM Tris + 0.01% Tween-20) was added, and the sample was mixed for 5 minutes on a Bioshake iQ shaker. The purified DNA was used directly for library preparation with the Ligation Sequencing Kit V14 (Oxford Nanopore Technologies; catalog number: SQK-LSK114) following the manufacturer’s protocol.

### Nanopore sequencing

Genomic DNA libraries were prepared using the Oxford Nanopore Ligation Sequencing Kit V14 (SQK-LSK114) on PromethION with R10.4.1 flow cells (FLO-PRO114M). Three µg of high-molecular-weight DNA were transferred to a 1.5 mL DNA LoBind tube and brought to 47 µl with nuclease-free water (Thermo Fisher Scientific, Cat. 10977015). DNA repair and end-prep were performed in a 0.2 mL PCR tube with the NEBNext Companion Module for Oxford Nanopore Ligation Sequencing (NEB, Cat. E7180L) following the manufacturer’s instructions, incubating at 20°C for 5 min and 65°C for 5 min, then cooling to 4°C on a thermocycler. The reaction was purified by SPRI using AMPure XP on a magnetic rack with two 80% ethanol washes and eluted in 61 µl nuclease-free water (Beckman Coulter, Cat. A63881). Sequencing adapters (LA) were ligated in ONT Ligation Buffer (LNB) with NEB Quick

T4 DNA Ligase from the module, followed by clean-up using ONT Long Fragment Buffer (LFB) to enrich fragments ≥3 kb and elution in 25 µl Elution Buffer (EB). Library yield was quantified fluorometrically (Qubit Fluorometer with dsDNA kit, Thermo Fisher Scientific, Cat. Q32853) and supported multiple reloads per flow cell. For loading, 32 µl of library were combined with Sequencing Buffer (SB) and Library Solution (LIS) to a final 200 µl mix. Flow cells were primed with Flow Cell Flush (FCF) and Flow Cell Tether (FCT), and 200 µl of the library were loaded per the manufacturer’s recommendations. Sequencing was run on a PromethION 48 under MinKNOW control for 72 h; the run was paused at 24 h, the flow cell washed (Flow Cell Wash Kit, EXP-WSH004) and reloaded, then resumed.

### Genome-in-a-Bottle benchmarking

Quality-passed nanopore sequencing reads from Oxford Nanopore Technologies were concatenated by sample and aligned using minimap2 with ONT-specific parameters to both GRCh37 (for structural variant analysis) and GRCh38 (for small variant calling) reference genomes. Small variants were called using DeepVariant v1.6.1 and benchmarked against GIAB truth sets using hap.py for trio concordance analysis. Structural variants (≥50 bp) were called using Sniffles2 (v2.2) and benchmarked against the GIAB HG002 truth set using Truvari v4.2.2. Variant calling performance was evaluated using precision, recall, and F1 metrics within GIAB-defined high-confidence regions.

### Variant and methylation review

Loci of interest (*GTF2I, KDM2A*) were inspected in IGV^24^ (version 2.19.2). CpG 5mC tracks were generated from native signals using the study pipeline and viewed alongside CpG-island annotations. For each locus, we report local coverage, variant-supporting read counts, approximate variant allele fraction, and the presence or absence of nearby structural variants within the inspected window.

### Exploratory methylation analysis

To illustrate the *KDM2A* episignature in native nanopore data, we visualized representative sites previously implicated in the signature (for example, CYP26C1 CpG255, RB1 CpG85, and HS3ST3B1 CpG267) and compared the proband qualitatively against ensilicated controls without *KDM2A* variants. Given the small sample size, these analyses are descriptive.

## Data Availability

All data produced in the present study are available upon reasonable request to the authors.

## Acknowledgements

We thank all research participants in the UDN. We would also like to thank members of the Ashley Lab, Wheeler Lab, the UDN, and the Stanford Center for Undiagnosed Diseases, who gave invaluable feedback and assistance throughout this project. Some figures were created in https://BioRender.com/t867ano

## Consortia

Members of the Undiagnosed Diseases Network: Alyssa A. Tran, Arjun Tarakad, Ashok Balasubramanyam, Brendan H. Lee, Carlos A. Bacino, Daryl A. Scott, Elaine Seto, Gary D. Clark, Hongzheng Dai, Hsiao-Tuan Chao, Ivan Chinn, James P. Orengo, Jill A. Rosenfeld, Kim Worley, Lindsay C. Burrage, Lisa T. Emrick, Lorraine Potocki, Monika Weisz Hubshman, Richard A. Lewis, Ronit Marom, Seema R. Lalani, Shamika Ketkar, Tiphanie P. Vogel, William J. Craigen, Jared Sninsky, Lauren Blieden, Sandesh Nagamani, Hugo J. Bellen, Michael F. Wangler, Oguz Kanca, Shinya Yamamoto, Christine M. Eng, Patricia A. Ward, Pengfei Liu, Adeline Vanderver, Cara Skraban, Edward Behrens, Gonench Kilich, Kathleen Sullivan, Kelly Hassey, Ramakrishnan Rajagopalan, Rebecca Ganetzky, Vishnu Cuddapah, Anna Raper, Daniel J. Rader, Giorgio Sirugo, Anne Slavotinek, Christopher Mayhew, Eneida Mendonca, Ziyuan Guo, Allyn McConkie-Rosell, Kelly Schoch, Mohamad Mikati, Nicole M. Walley, Rebecca C. Spillmann, Vandana Shashi, Alan H. Beggs, Calum A. MacRae, David A. Sweetser, Deepak A. Rao, Edwin K. Silverman, Elizabeth L. Fieg, Frances High, Gerard T. Berry, Ingrid A. Holm, J. Carl Pallais, Joan M. Stoler, Joseph Loscalzo, Lance H. Rodan, Laurel A. Cobban, Lauren C. Briere, Matthew Coggins, Melissa Walker, Richard L. Maas, Susan Korrick, Jessica Douglas, Cecilia Esteves, Emily Glanton, Isaac S. Kohane, Kimberly LeBlanc, Shamil R. Sunyaev, Shilpa N. Kobren, Brett H. Graham, Erin Conboy, Francesco Vetrini, Kayla M. Treat, Khurram Liaqat, Lili Mantcheva, Stephanie M. Ware, Kathleen Page, Paul Auwaerter, Yuka Manabe, Carlos A. Pardo-Villamizar, Julie Hoover-Fong, Philip Dane Witmer, Winston Timp, Matthew Robinson, Zackary Dov Berger, Elizabeth Wohler, Nara Sobreira, Arian Nouraee, Carlos Prada, Erica Davis, Kai Lee Yap, Kelly Regan-Fendt, María Paula Silva, Patrick McMullen, Breanna Mitchell, Brendan C. Lanpher, Devin Oglesbee, Eric Klee, Filippo Pinto e Vairo, Ian R. Lanza, Kahlen Darr, Lindsay Mulvihill, Lisa Schimmenti, Queenie Tan, Surendra Dasari, Abdul Elkadri, Brett Bordini, Donald Basel, James Verbsky, Julie McCarrier, Michael Muriello, Michael T. Zimmermann, Adriana Rebelo, Carson A. Smith, Deborah Barbouth, Guney Bademci, Joanna M. Gonzalez, Kumarie Latchman, LéShon Peart, Mustafa Tekin, Nicholas Borja, Stephan Zuchner, Stephanie Bivona, Willa Thorson, Herman Taylor, Rakale C. Quarells, Ayuko Iverson, Bruce Gelb, Charlotte Cunningham-Rundles, Eric Gayle, Joanna Jen, Louise Bier, Mafalda Barbosa, Manisha Balwani, Mariya Shadrina, Rachel Evard, Saskia Shuman, Susan Shin, Vaidehi Jobanputra, Andrea Gropman, Barbara N. Pusey Swerdzewski, Camilo Toro, Colleen E. Wahl, Donna Novacic, Ellen F. Macnamara, John J. Mulvihill, Maria T. Acosta, Precilla D’Souza, Valerie V. Maduro, Ben Afzali, Ben Solomon, Cynthia J. Tifft, David R. Adams, Elizabeth A. Burke, Francis Rossignol, Heidi Wood, Jiayu Fu, Joie Davis, Leoyklang Petcharet, Lynne A. Wolfe, Margaret Delgado, Marie Morimoto, Marla Sabaii, MayChristine V. Malicdan, Neil Hanchard, Orpa Jean-Marie, Wendy Introne, William A. Gahl, Yan Huang, Andrew Stergachis, Danny E. Miller, Elisabeth Rosenthal, Elizabeth Blue, Elsa Balton, Emily Shelkowitz, Eric Allenspach, Fuki M. Hisama, Gail P. Jarvik, Ghayda Mirzaa, Ian Glass, Kathleen A. Leppig, Katrina Dipple, Mark Wener, Martha Horike-Pyne, Michael Bamshad, Peter Byers, Runjun Kumar, Seth Perlman, Sirisak Chanprasert, Virginia Sybert, Wendy Raskind, Nitsuh K. Dargie, Chun-Hung Chan, Dr. Francisco Bustos velasq, Isum Ward, Jason Schend, Jennifer Morgan, Megan Bell, Miranda Leitheiser, Mohamad Saifeddine, Paul Berger, Rachel Li, Taylor Beagle, Alexander Miller, Beatriz Anguiano, Beth A. Martin, Brianna Tucker, Chloe M. Reuter, Devon Bonner, Elijah Kravets, Hector Rodrigo Mendez, Holly K. Tabor, Jacinda B. Sampson, Jason Hom, Jennefer N. Kohler, Jennifer Schymick, John E. Gorzynski, Jonathan A. Bernstein, Kevin S. Smith, Laura Keehan, Laurens Wiel, Matthew T. Wheeler, Meghan C. Halley, Mia Levanto, Page C. Goddard, Paul G. Fisher, Rachel A. Ungar, Raquel L. Alvarez, Sara Emami, Shruti Marwaha, Stephen B Montgomery, Suha Bachir, Tanner D Jensen, Taylor Maurer, Terra R. Coakley, Euan A. Ashley, Anna Hurst, Brandon M Wilk, Bruce Korf, Elizabeth A Worthey, Kaitlin Callaway, Martin Rodriguez, Pongtawat Lertwilaiwittaya, Reaford Blackburn, Tammi Skelton, Tarun KK Mamidi, Teneasha Washington, Andrew B. Crouse, Jordan Whitlock, Mariko Nakano-Okuno, Matthew Might, William E. Byrd, Albert R. La Spada, Changrui Xiao, Elizabeth C. Chao, Eric Vilain, Jose Abdenur, Kirsten Blanco, Maija-Rikka Steenari, Rebekah Barrick, Richard Chang, Sanaz Attaripour, Suzanne Sandmeyer, Tahseen Mozaffar, Alden Huang, Andres Vargas, Bianca E. Russell, Brent L. Fogel, Esteban C. Dell’Angelica, George Carvalho, Julian A. Martínez-Agosto, Layal F. Abi Farraj, Manish J. Butte, Martin G. Martin, Naghmeh Dorrani, Neil H. Parker, Rosario I. Corona, Stanley F. Nelson, Yigit Karasozen, Dana Sayer, Jennifer Tousseau, Aaron Quinlan, Alistair Ward, Ashley Andrews, Corrine K. Welt, Dave Viskochil, Erin E. Baldwin, John Carey, Justin Alvey, Lorenzo Botto, Nicola Longo, Paolo Moretti, Rebecca Overbury, Russell Butterfield, Steven Boyden, Thomas J. Nicholas, Matt Velinder, Gabor Marth, Pinar Bayrak-Toydemir, Rong Mao, Monte Westerfield, John A. Phillips III, Kimberly Ezell, Lynette Rives, Rizwan Hamid, Alyson Krokosky, Ashley McMinn, Cathy Shyr, Eric Gamazon, Joy D. Cogan, Lakshitha Perera, Lisa Bastarache, Mary Koziura, Thomas Cassini, Alex Paul, Dana Kiley, Daniel Wegner, Erin McRoy, Jennifer Wambach, Kathy Sisco, Patricia Dickson, F. Sessions Cole, Dustin Baldridge, Jimann Shin, Lilianna Solnica-Krezel, Stephen C. Pak, Timothy Schedl, Allen Bale, Carol Oladele, Caroline Hendry, Emily Wang, Hua Xu, Hui Zhang, Lauren Jeffries, María José Ortuño Romero, Mark Gerstein, Michele Spencer-Manzon, Monkol Lek, Nada Derar, Odelya Kaufman, Shrikant Mane, Teodoro Jerves Serrano, Vasilis Vasiliou, Winston Halstead, Yong-Hui Jiang

## Ethics Declaration

Ethical and research approvals for participants in the Undiagnosed Diseases Network were obtained from the Stanford University Institutional Review Board (protocols 47026) and the National Human Genome Research Institute Institutional Review Board (protocol 15-HG-0130). All participants provided written informed consent.

