## Supplementary Information for "Ensilication preserves high-molecular weight native DNA for clinical long-read sequencing"

**Supplementary Section 1. Ensilication provides robust protection for ultra-long DNA under routine handling and accelerated weathering.**

For traditionally stored samples at –80 °C, repeated freeze-thaw cycles are unavoidable during normal laboratory use, which can result in fragmentation and eventual loss of DNA ^1,2^. Initial fragment analysis showed similar DNA integrity profiles between storage and handling conditions: freeze-thaw cycles for –80 °C samples and hydration-dehydration cycles with vortexing for ensilicated samples (**Supplementary Fig. 1**). However, when we employed nanopore sequencing, distinct differences emerged in DNA preservation profiles. N50 analysis revealed that repeated freeze-thaw cycles impacted DNA integrity in –80 °C samples across all three reference genomes (**Supplementary Fig. 1a**). HG002 showed a 2.0 kb reduction in N50 (9.60 to 7.60 kb) after freeze-thaw cycles, while ensilicated samples demonstrated better resilience with only a 0.8 kb decrease (9.10 to 8.30 kb) after multiple rounds of hydration-dehydration cycles with vortexing to ensure a well-dispersed colloidal solution. Similarly, HG003 samples subjected to freeze-thaw cycles showed a marked decline in N50 (9.00 to 6.60 kb), while ensilicated samples maintained better integrity (8.00 to 7.80 kb) despite hydration stress. This pattern was consistent in HG004, where freeze-thaw cycles resulted in a 2.8 kb decrease (10.00 to 7.20 kb), compared to a more modest 2.0 kb reduction (10.60 to 8.60 kb) in ensilicated samples under hydration-dehydration stress.

To evaluate long-term stability at ambient temperature, we employed an accelerating weathering protocol based on established approaches from materials science^3,4^ and the pharmaceutical industry^5,6^. These protocols leverage the predictable relationship between temperature and reaction kinetics, though they assume consistent degradation mechanisms across temperatures and constant environmental conditions besides temperature. Using the temperature coefficient (Q_10_), a measure describing how reaction rates change with temperature^7,8^, and a previously established Q_10_ of 6.9 for DNA degradation from a similar encapsulation material for ensilicated samples^9^, we calculated that exposure to 55°C for 21 days would simulate approximately 10,000 days of storage at a room temperature of 23°C.

Using this accelerated weathering protocol, we compared ensilicated DNA against dried genomic, desalted DNA. Drying DNA in sealed containers has been widely explored as it removes water molecules that contribute to hydrolytic damage, potentially enabling preservation without specialized cold chain infrastructure^10,11^. To evaluate the protective effects of ensilication versus simple DNA drying, we subjected both sample types to accelerated weathering conditions at 70% relative humidity. Fragment size distribution analysis revealed distinct degradation patterns between the two preservation approaches (Fig. 1b). While both storage approaches initially displayed characteristic peaks around 10^5^ bp at day 0, ensilicated samples demonstrated resistance to these harsh conditions, maintaining a more stable size distribution profile particularly in the critical 10^4^–10^5^ bp range. In contrast, dried DNA samples showed progressive fragmentation, with a pronounced shift toward smaller fragment sizes after 21 days at 55 °C and 70% relative humidity.

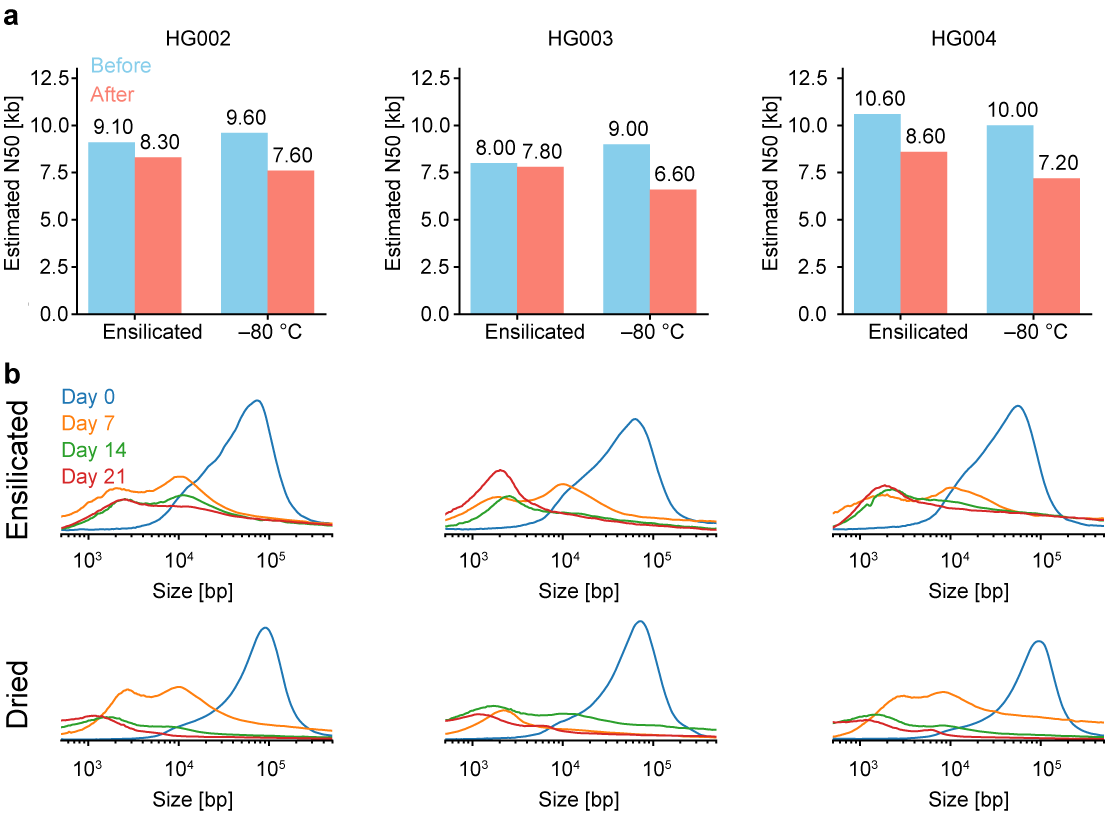

**Supplementary Figure 1 | Comparative stability of DNA preservation methods under freeze-thaw cycles and accelerated weathering. a,** N50 analyses of ensilicated samples after 19 cycles of hydration-dehydration with vortexing and frozen samples after 19 freeze-thaw cycles. **b,** Comparison of electropherograms between ensilicated and dried samples under accelerated weathering conditions.
